# Characteristics of lymphocyte subsets and their predicting values for the severity of COVID-19 patients

**DOI:** 10.1101/2020.05.01.20086421

**Authors:** Jingrong Wang, Xingqi Dong, Boting Zhang, Xinping Yang, Zhi Li, Xicheng Wang, Shuguang Zuo

**Author notes:** These authors contributed equally to this article (Jingrong Wang, Xingqi Dong). Correspondences (X.WANG); (S.ZUO).

## Abstract

Severe COVID-19 patients showed worse clinical outcomes compared to mild and moderate patients. However, effective indicators are still lacking to predict the severity of the disease. In the present study, we retrospectively analyzed the clinical and laboratory data of 16 COVID-19 patients and found that the absolute counts of three T-cells (CD3^+^, CD4^+^, and CD8^+^) were significantly lower in the moderate and severe patients than those in mild patients and were significantly lower in severe patients than in moderate patients on admission. With the recovery of the COVID-19, serum levels of inflammatory biomarkers (CRP, PCT, and IL6) of moderate and severe patients gradually decreased. In contrast, the counts of lymphocytes and their subsets including CD3^+^, CD4^+^, and CD8^+^ T cells gradually increased in severe patients, and eventually showed comparable levels with moderate patients. ROC analysis showed that the counts of CD3^+^, CD4^+^, and CD8^+^ T-cells with AUC > 0.9 have potential values for predicting the severity of COVID-19 patients. In conclusion, the reduction of CD3^+^, CD4^+^, and CD8^+^ T-cells is related to the severity of COVID-19 and dynamic detection of these T-lymphocyte subsets may help predict the outcome of the patients.

## Introduction

In December 2019, a novel coronavirus (provisionally called 2019-nCoV) infected pneumonia occurred in Wuhan, Hubei Province, China[1]. Phylogenic analysis indicated that 2019-nCoV is a positive-chain RNA enveloped beta genus coronavirus, and closely related to Severe Acute Respiratory Syndrome Coronavirus (SARS-CoV) and Middle East respiratory syndrome Coronavirus (MERS-CoV)[2, 3]. The disease caused by 2019-nCoV was recently named by the World Health Organization (WHO) as Coronavirus Disease 2019 (COVID-19)[4]. Shortly after WHO announced the official name of the disease, 2019-nCoV is officially named as Severe Acute Respiratory Syndrome Coronavirus 2 (SARS-CoV-2) by the International Committee on Taxonomy of Viruses (ICTV)[5]. According to the daily report of the WHO, by February 25,2020, the epidemic of SARS-CoV-2 has caused 80,239 confirmed cases, of which 77,780 were confirmed in China, 2,666 were dead, and 2,459 were confirmed in 33 countries outside China, 34 patients died[6]. COVID-19 caused by SARS-CoV-2 has posed a serious threat to global public health.

SARS-CoV-2 can cause different clinical types (severity) of COVID-19. Mild and some of the moderate patients can heal themselves, but other moderate patients will develop into severe patients. The severe patients with underlying diseases often develop organ dysfunction (such as shock, acute respiratory distress syndrome (ARDS), acute heart injury, and acute renal failure), and some critical patients require admission to the intensive care unit (ICU). Therefore, it is very imperative to find indicators that can predict the severity of the disease.

COVID-19 patients are commonly accompanied by an increase of inflammatory biomarkers [e.g., C-reactive protein (CRP), procalcitonin (PCT), and interleukin 6 (IL6)], leukopenia and lymphopenia[7, 8], which is more pronounced in severe patients. However, the status of the lymphocyte subpopulations especially the T cell subsets in these patients are not well understood. Furthermore, the dynamic changes of the previously mentioned inflammatory biomarkers and the lymphocyte subpopulations in COVID-19 patients remain unclear. With this in mind, we retrospectively analyzed three commonly used inflammatory biomarkers (CRP, PCT, and IL6), T-cell subsets in the peripheral blood as well as the clinical data of 16 SARS-CoV-2 infected patients to identify potential predictors regarding the severity of the COVID-19 patients.

## Methods

### Data collection

The study was approved by the Ethics Committee of Yunnan Provincial Hospital of Infectious Disease, AIDS Care Center. Oral consent was obtained from patients. The 16 laboratory-confirmed COVID-19 patients at Yunnan Provincial Hospital of Infectious Disease from January 17 to February 8, 2020, were enrolled in this retrospective study. All medical record information, including epidemiological, clinical, radiological, laboratory and treatment data, was obtained through data collection forms of electronic medical records. All data reviewed by a team of trained physicians.

### Laboratory examination

Throat swab samples were collected from patients suspected of having SARS-CoV-2 infection and maintained in a collection tube with the virus-preservation solution. After collection, total RNA was extracted within 2 hours using the virus RNA isolation kit (DAAN Gene Co., Ltd. of Sun Yat-sen University, Guangzhou, China). The quantitative reverse transcription PCR (RT-qPCR) assay was performed using a SARS-CoV-2 nucleic acid detection kit according to the manufacturer’s protocol (Shanghai GeneoDx Biotechnology Co., Ltd., and Shanghai BioGerm Medica Biotechnology Co., Ltd., Shanghai, China). Lymphocyte subsets were detected by flow cytometry using a 4-color BD Multitest™ CD3/CD8/CD45/CD4 kit (BD Biosciences, USA) according to the manufacturer’s protocol. Flow cytometry was performed on FACSCalibur (BD Biosciences, USA).

### Clinical classification of the COVID-19 patients

Clinical classification of the COVID-19 patients was performed according to the “Guidelines of the Diagnosis and Treatment of COVID-19 Pneumonia (Version 5)” published by the National Health Commission of China. Mild patients: the clinical symptoms of patients are mild, and there is no pneumonia on CT imaging. Moderate patients: patients with fever and respiratory symptoms, CT imaging show pneumonia. Severe patients should meet at least one of the following conditions: shortness of breath and the respiratory rate (RR) is greater than or equal to 30 times/min; Finger oxygen saturation at resting is less than or equal to 93%; arterial oxygen partial pressure to fractional inspired oxygen (PaO_2_/FiO_2_ ratio) is less than or equal to 300. Critical patients should meet at least one of the following conditions: patients have respiratory failure and need mechanical ventilation; Patients have shock; Patients with other organ failure and require monitoring in the ICU.

### Statistical Analysis

All statistical analyses were performed using R (version 3.5.1). Categorical variables were described using frequency or percentage, and continuous variables were described as mean, median, and quartile intervals. When the data were Gaussian distributed, the mean of continuous variables was compared using ANOVA, and the differences between the 3 groups were tested by Tukey’s HSD test. When the data were not Gaussian distributed, the median of continuous variables was compared using non-parametric analysis and the significance is tested by the Kruskal Wallis test. Significance between the two groups is tested by the Wilcoxon signed-rank test. Correlation analysis was performed using Spearman’s method. The ggplot2 package (Version 3.2.1, http://ggplot2.tidyverse.org, https://github.com/tidyverse/ggplot2) was used to visualize data with R. In all statistical analyses, *P* < 0.05 was considered statistically significant. Receiver operating characteristic (ROC) curves were used to assess the accuracy of variables predicting severely ill patients. A pROC package (Version 1.16.1, http://expasy.org/tools/pROC) was used to visualize the ROC curve and calculate the area under the curve (AUC).

## Results

### Clinical characteristics of COVID-19 patients

All admitted patients were confirmed by RT-qPCR measurement of SARS-CoV-2 RNA in throat swab samples. The 16 COVID-19 patients were divided into three groups according to previously mentioned Guidelines, including 5 mild, 7 moderate, and 4 severe cases [Oxygen saturation decreased (≤90%)]. The median age was 49 years (3-69 years) in total patients, of which the median age of the severe group (61, 49-69 years) was older than that of the moderate (50, 32-57 years) and mild group (22, 3-40 years). The 16 patients included 9 males (3 in each of groups) and 7 females (2 were mild, 4 were moderate, and 1 was severe). Except for one patient who is a native of Kunming (with a history of tourism in Wuhan), the remaining patients are from Wuhan and have no history of contact with the seafood market. Four patients were family clustered, of which three patients were mild and one patient was moderate. The median time from the onset of symptoms to admission was 6 days (2 days for mild patients, 7 days for moderate patients, and 7.5 days for severe patients). Three patients had hypertension (one in the mild group, two in the severe group), and three patients had diabetes (one in the moderate group, two in the severe group). Twelve patients had a fever, of which the mild, moderate and severe groups included 3, 5, and 4 patients, respectively. Of the 16 patients, 7 had a cough, 6 had sputum production, 2 had chest tightness, 2 had shortness of breath, 6 had fatigue, 1 had a headache, 1 had a sore throat, and 2 had body aches. CT imaging of 6 patients was normal, and the other 10 patients were abnormal. Patients with abnormal CT imaging showed abnormalities in the subpleural, pulmonary base, and bronchoalveolar segments, which is consistent with atypical upper respiratory symptoms. Although the CT scan showed abnormal lesions close to the pleura, no pleural fluid was found in all patients, indicating that the pleura was not affected, which is consistent with the absence of chest pain in all patients. Three patients were treated with antibiotics (piracillin and sulbactam, moxifloxacin, and meropenem). All patients received antiviral therapy with inhaled IFN-α for 7 days. Fourteen patients received KALETRA [(lopinavir/ritonavir), BID, 500 mg/day], of which 12 patients completed a 14-day course, and one severe patient was discontinued because of vomiting after 2 doses of medication, and another severe patient was stopped due to hepatic dysfunction. Five patients received immunoglobulin (400 mg/kg/day), of which one was a moderate patient and four were severe patients. Of the four severe patients, one patient received a 4-day course of glucocorticoids (prednisone, 80 mg/kg/day). As of February 24, 2020, 13 of the 16 patients had SARS-CoV-2 RNA converted to negative, and one patient in each group had not converted to negative RNA. The median days from onset to virus eradication was 21 days, 17 days for mild patients, 24 days for moderate patients, and 21 days for severe patients.

### The Counts of lymphocyte subsets were associated with the severity of the COVID-19 patients

To find laboratory indicators associated with the severity of the disease at admission, we compared the differences of CRP, PCT, IL6, WBC, neutrophil, lymphocyte (LC), and T-lymphocyte subsets (CD3^+^, CD4^+^, and CD8^+^ T) cells among the 3 clinical types of COVID-19 patients. As shown in **Figure 1**, Except for PCT, serum levels of CRP and IL6 were significantly higher in the severe group than in the mild and moderate group (all P<0.05). LC counts (P=0.02), CD3^+^ T-cell counts and ratio (P=0.019, P=0.02), CD4^+^ T-cell counts (P=0.02), CD8^+^ T-cell counts and ratio (P=0.019, P=0.02) were significantly lower in the severe group compared to the mild group. In contrast, NLR (neutrophils to lymphocyte ratio) were significantly higher in the moderate group than in the mild group (P=0.015). Of note, only CD3^+^ T-cell counts (P=0.029), CD4^+^ T-cell counts (P=0.047) and CD8^+^ T-cell counts (P=0.047) were significantly lower in the severe group than in the moderate group. Except for CD3^+^, CD4^+^ and CD8^+^ T-cells counts, no differences in these indicators were observed between the severe and the moderate group. These data suggest that the decreased CD3^+^, CD4^+^ and CD8^+^ T-cell counts have the potential value to predict the severity of COVID-19 patients.

**Figure 1.**
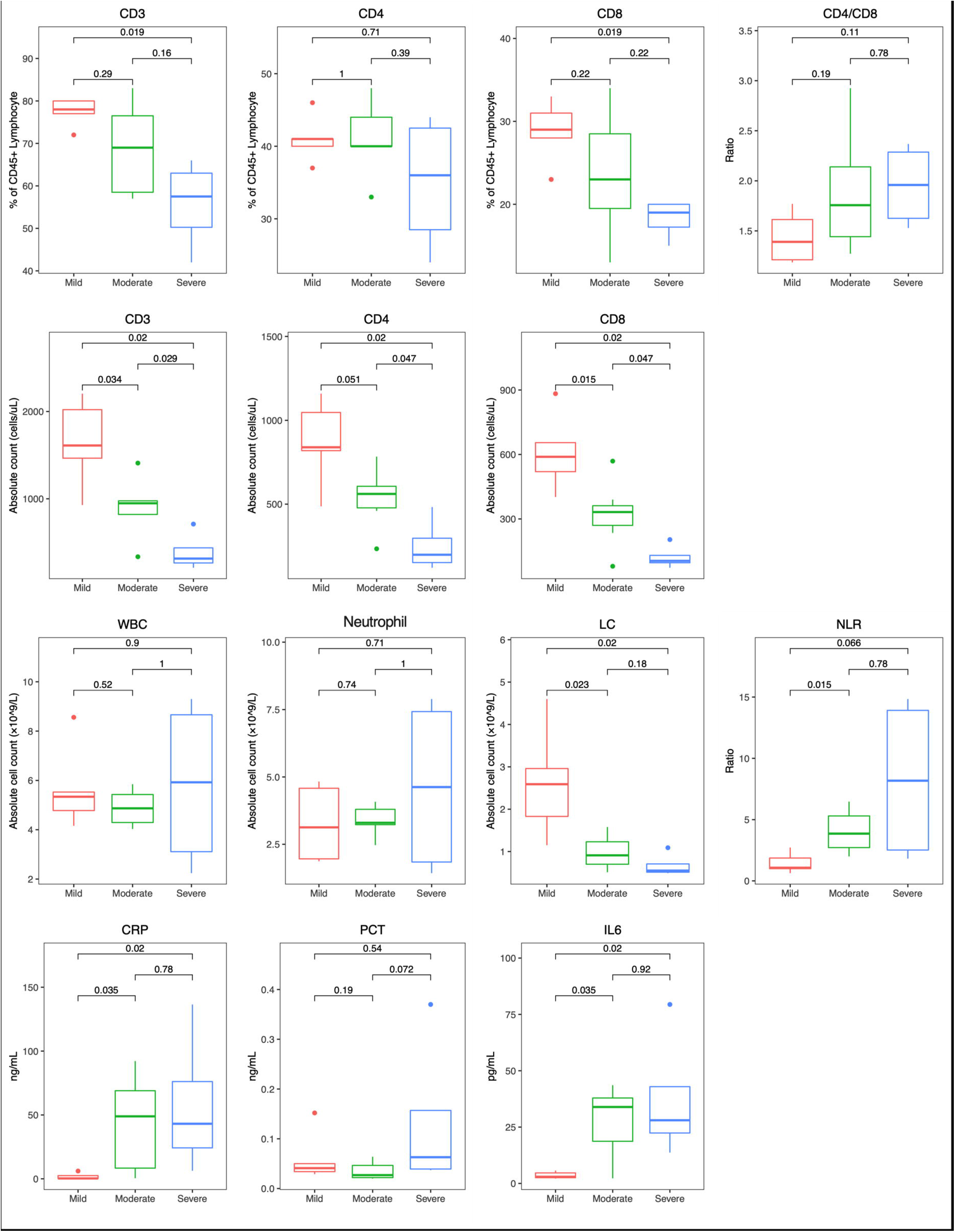
Compare the indicators in the different type (severity) of COVID-19 patients on admission. The lines in the middle of the boxes represent the median, boxes represent 25th and 75th percentiles and whiskers represent 5th and 95th percentiles. Significance between different groups is tested by the Kruskal Wallis test. A p-value of less than 0.05 is statistically significant.

### Dynamic analysis of the indicators in moderate and the severe COVID-19 patients

We analyzed the dynamic changes of CRP, PCT, IL6, WBC, Neutrophil, LC in moderate and severe groups. As shown in **Figure 2**, CRP levels gradually decreased in both moderate and severe groups. PCT levels continued to be lower from 6 to 21 days in the moderate group, whereas it was initially higher and then gradually decreased in the severe group. In the severe group, IL6 levels peaked at days 8-11 but then continued to decline. In the moderate group, IL6 showed a downward trend in fluctuations. Except for days 12-14, there was no significant difference in IL6 levels between the two groups. At days 6-8 after disease onset, the LC counts in the moderate group were higher than that of the severe group, but there was no statistical difference. In the following days, the LC counts of the moderate group gradually reached their highest levels and kept close to normal levels (1.5 × 10^9^ cells/L). The LC counts in the severe group gradually decreased, reached its lowest value on days 12-14, and then gradually increased. On days 18-20, LC counts in the severe group reached a comparable level to the moderate group. However, there were statistically significant differences in LC counts between the two groups over three consecutive time periods (P<0.05). There were no significant differences in WBC counts, Neutrophil counts, and NLRs between the two groups throughout the course of the disease.

**Figure 2.**
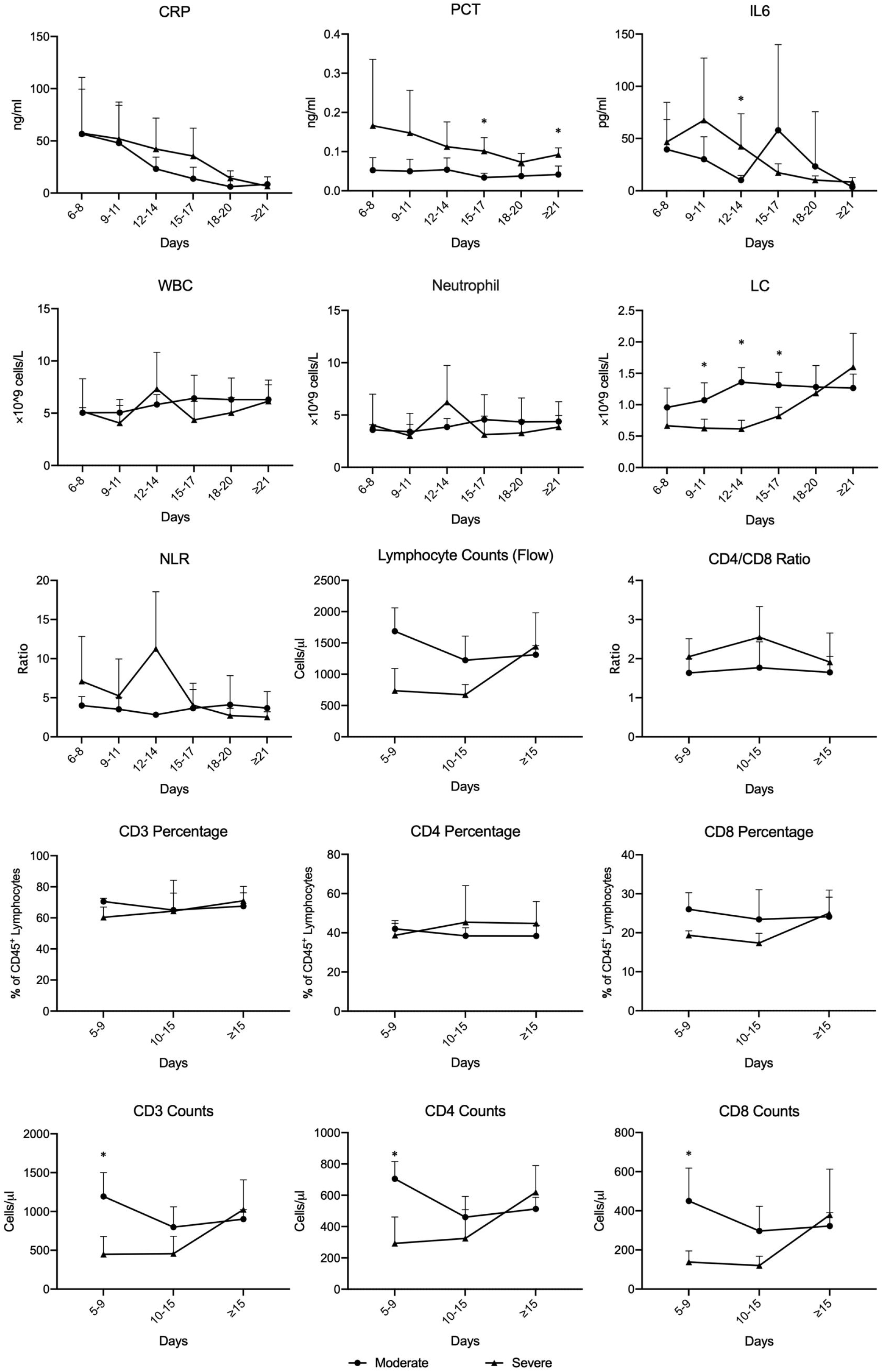
Dynamic analysis of indicators in moderate and the severe COVID-19 patients. The lines represent mean, error bar represents SD. Significance between different groups is tested by the Wilcoxon signed-rank test. Asterisk (*) indicates significant (P<0.05) differences between the two groups in various time points.

Next, we analyzed the dynamic changes of CD3^+^, CD4^+^ and CD8^+^ T-cells in the moderate and the severe COVID-19 patients. As shown in **Figure 2**, the CD3^+^, CD4^+^ and CD8^+^ T-cell counts in the moderate group were significantly higher than that of the moderate group initially (P<0.05). However, the counts of lymphocytes and their subsets including CD3^+^, CD4^+^, and CD8^+^ T-cells in the severe group gradually increased. With the disease recovery, all these cells reached a comparable level to the moderate group. There were no significant differences in the percentage of CD3^+^, CD4^+^ and CD8^+^ T-cells, CD4/CD8 ratio between the two groups throughout the course of the disease.

To investigate the association between these indicators, we performed Spearman’s correlation analysis. As shown in **Figure 3**, LC counts were negatively correlated with the majority of variables (R< −0.3, P<0.05) except for WBC and Neutrophil counts. In contrast, NLR was positively correlated with all the other variables (R> 0.3, P<0.05) except for LC (R< −0.3). Levels of CRP, PCT and IL6 displayed a positive correlation with each other (R> 0.3, P<0.05). Of note, IL6 was positively correlated with other variables (R> 0.3, P<0.05), but negatively correlated with LC counts (R< −0.3, P<0.05). Moreover, counts of lymphocytes and their subsets including CD3^+^, CD4^+^, and CD8^+^ T-cells displayed a positive correlation with each other (R> 0.9, P<0.05).

**Figure 3.**
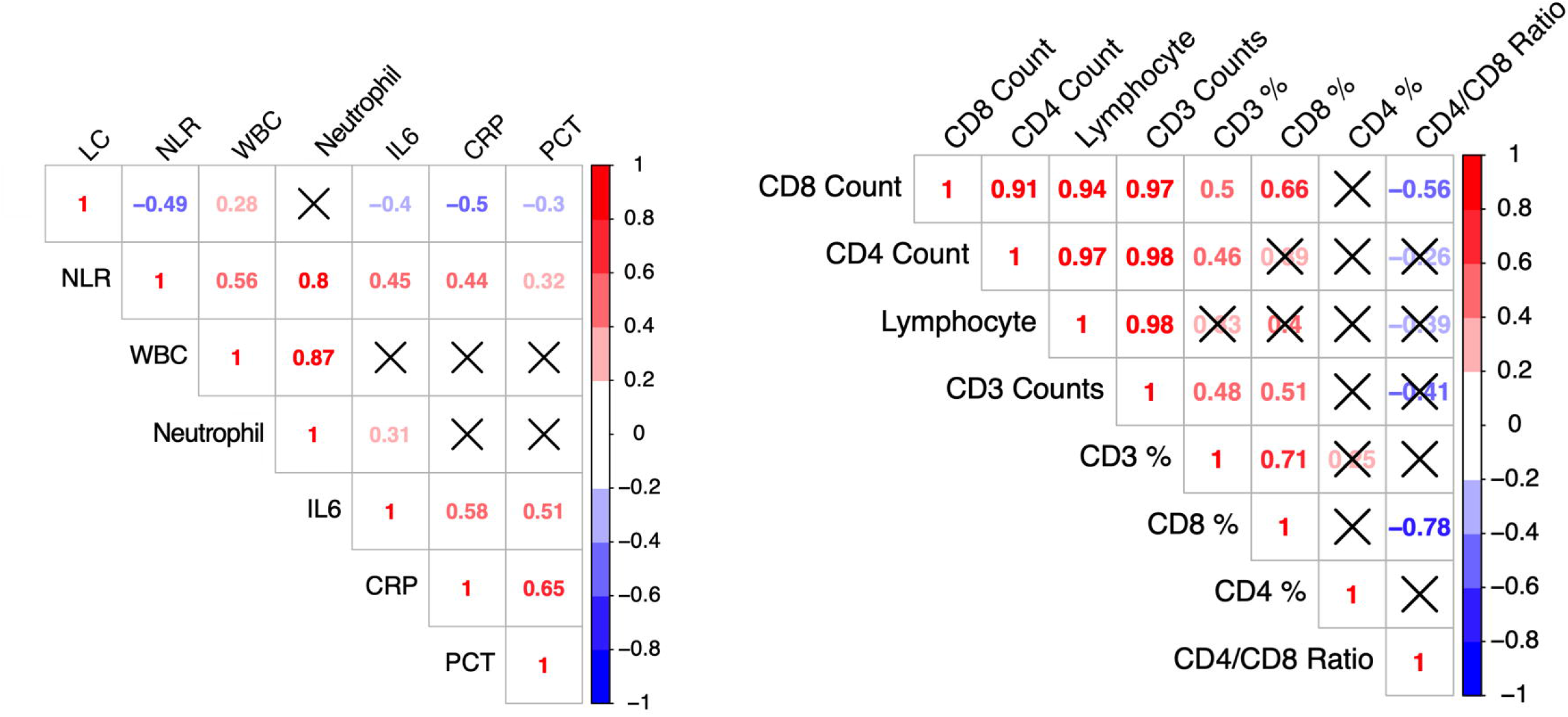
Correlation analysis between indicators. The correlation coefficient (R) was calculated using Spearman’s correlation method. R between 0.3 and 0.7 (−0.3 and −0.7) indicate a moderate positive (negative) correlation; R between 0.7 and 1 (−0.7 and −1) indicate a strong positive (negative) correlation. Mark “X” indicates no significant difference (P>0.05).

### Counts of T cells are potential predictors for severe COVID-19 patients

To investigate the potential predictive value of these parameters, a receiver operating characteristic (ROC) analysis was performed and the area under ROC curve (AUC) was calculated. As shown in **Figure 4**, four variables (including the counts of lymphocyte, CD3^+^, CD4^+^, and CD8^+^ T-cells) had an AUC of greater than 0.9, whereas other variables had an AUC of lesser than 0.9. Furthermore, the cutoff values of the four parameters with higher AUC were calculated from the ROC curves, with a value of 0.855 x 10^9^ cell/L for lymphocyte, 582 cells/μL for CD3^+^ T-cells, 347 cells/μL for CD4^+^ T-cell and 171 cells/μL for CD8^+^ T-cells. Both CD3^+^ and CD4^+^ T-cells showed the highest AUC which suggesting a more powerful prediction performance for severe COVID-19 patients.

**Figure 4.**
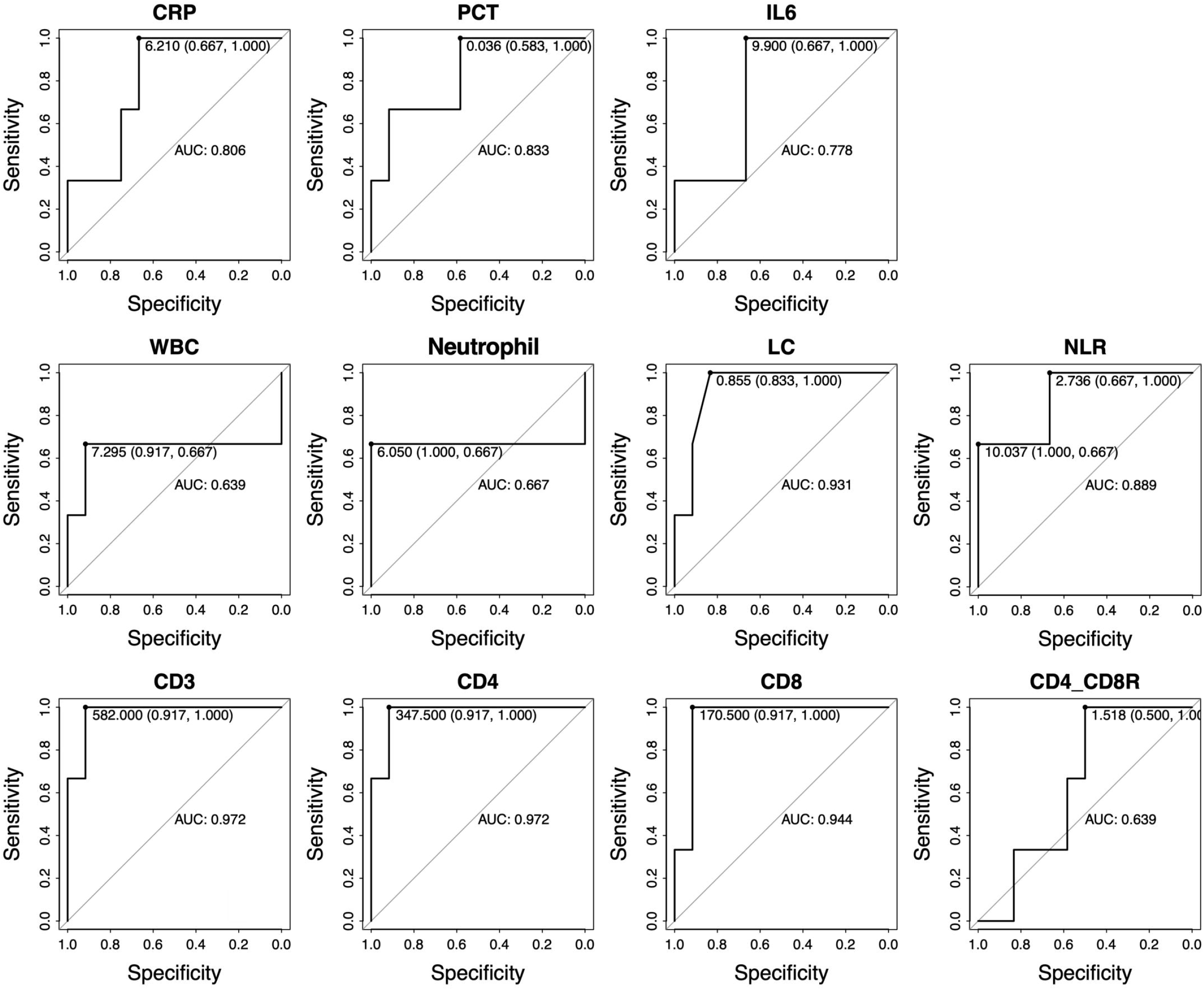
Receiver operator characteristic (ROC) analysis of the biomarker performance. The best cutoff values of the parameters with higher areas under the curve (AUC) were calculated from the ROC curves.

## Discussion

Previous reports showed that the majority (63%-70%) of COVID-19 patients on admission have lymphopenia[9, 10], and nonsurvivors develop more severe lymphopenia throughout the course of the disease[10]. A case report also confirmed this phenomenon, in which the nonsurvivor’s lymphocytes continued to decrease[11]. These papers suggest that lymphopenia might be a common feature in the COVID-19 patients and the lymphocyte status might be a critical factor related to disease severity and mortality. In addition to confirming the previously reported lymphopenia in the COVID-19 patients, we found that the counts of lymphocytes as well as their subsets CD3^+^, CD4^+^, and CD8^+^ T-cells are significantly decreased in severe patients compare to mild and moderated patients on admission. We also found that the lymphocytes especially the T-lymphocyte subsets gradually increased in severe patients with the recovery of COVID-19 patients, suggesting that the recovery of T-lymphocytes is associated with a favorable outcome in severe COVID-19 patients.

Previous studies have demonstrated a correlation between T-cell counts and the severity of SARS-CoV and MERS-CoV infected patients[12], as well as explored the possible mechanisms[13, 14]. Both SARS-CoV and MERS-CoV were found to be able to directedly infect the T cells and induced the lymphocyte apoptosis[13, 14]. Other studies have attributed the activation of the hypothalamus-pituitary-adrenal (HPA) axis under stress, which leads to excessive cortisol that causes apoptosis and decreases of lymphocytes [15]. This phenomenon is also seen in other patients with severe infections, such as sepsis[16]. Currently, there is no public evidence that steroid therapy can improve the incidence or mortality of SARS[17], but it can further aggravate the severity of lymphopenia[15]. Because lymphopenia is related to the severity of COVID-19 patients, caution should be exercised in using glucocorticoids in these patients. Even if low-dose glucocorticoids are needed to reduce pulmonary edema in patients with COVID-19, cortisol concentrations and the status of lymphocyte subsets in peripheral blood should be measured. It is unclear whether the T-cell reduction is caused by SARS-CoV-2 direct infection of T-cells or by indirect effects caused by systemic stress responses (such as HPA dysfunction). Further research is needed to explore the corresponding mechanism in detail.

Effective predictors can help doctors provide appropriate supportive care for patients with severe COVID-19. In this study, we found that four variables including lymphocyte, CD3^+^, CD4^+^, and CD8^+^ T-cells showed the potential value to predict severe patients. Both CD3^+^ and CD4^+^ T-cells showed the highest AUC (0.972) which suggested a more powerful predictive performance. A recent study reported that the NLR was a powerful prognostic factor for severe COVID-19 patients [18]. In their study, patients aged 50 years or older and NLR equal to or greater than 3.13 may serve as a high-risk factor for COVID-19 patients. In our study, we found that NLRs for all severe patients all exceeded 5, and NLRs for moderate patients ranged from 3 to 4. Moreover, the AUC of NLR for predicting severe patients is less than 0.9, indicating poor predictive performance. Another study showed that N8R (neutrophils to CD8^+^ T-cell ratio) has better performance with a higher AUC value than NLR in the ROC curve analysis, and may serve as a more powerful factor than NLR for predicting the severity in COVID-19 patients[19]. However, neutrophils are susceptible to many factors, and its ratio to CD8^+^ T cells will mask the actual reduction of T cells. Therefore, we believe that using direct counting of T cells may more accurately reflect the patient’s immune status and thus have better predictive performance.

Previous studies have shown that CD4^+^ T-cell counts are the main prognostic factor for AIDS patients [20]. When the counts of CD4^+^ T cells are below 200 cells/μL, patients are prone to secondary infections and tumors. HIV mainly infects CD4^+^ T cells and causes them to decline. However, in the early stages of HIV infection, CD8^+^ T cells increase reactively[21]. In this study, we found that the CD4^+^ and CD8^+^ T cells of COVID-19 patients decreased in a fixed ratio, which is different from HIV-infected patients. In addition, we found that the cutoff values for CD4^+^ and CD8^+^ T cells are 347 and 171 cells/μL, respectively. Due to the small sample size of our study, there may be some deviation in the cutoff values. A larger sample size study is needed to determine the best cutoff values. Nonetheless, the cutoff values we obtained are informative.

CRP, PCT, IL6 are commonly used inflammatory indicator for bacterial infections [22]. CRP is an acute-phase protein that is an extremely sensitive but non-specific biomarker in many kinds of systemic inflammatory processes, including bacterial and viral infections[23]. PCT has been proposed to be a reliable marker for distinguishing bacterial infections from other types of infections [24]. IL6 is always used for monitoring early states of infectious because of its fast serum peak[25]. In patients with COVID-19, about half of the patients had elevated concentrations of CRP (>10 mg/L)[26, 27]. Levels of IL6 showed sustained increases in severe patients compared to the mild patients[19]. In this study, we found that levels of CRP and IL6 were significantly higher in the moderate and severe patients than in mild patients on admission. With the recovery of COVID-19 patients, the elevated CRP and IL6 gradually decreased to normal levels. However, there was no difference in CRP, PCT, and IL6 levels between moderate and severe patients. ROC curve analysis also indicated that all of these indicators could not predict the severity of the disease. In SARS patients, the IL6 concentration was increased and positively correlated with the severity of the disease[28]. Although we found that IL6 does not predict the severity of COVID-19 patients, it does not mean that testing for IL6 is useless. It is well known that IL6 plays a key role in cytokine release syndrome (CRS) in chimeric antigen receptor (CAR) T-cell therapy of leukemia, as CRS patients can be found to have high levels of IL-6[29]. IL6 receptor (IL6R) antibody (e.g. Tocilizumab) can effectively control the CRS caused by CAR-T treatment[30]. Cytokine storms can usually be observed in patients with severe COVID-19[9]. Of course, the mechanism of cytokine storm caused by SAR-COV-2 may be different from CAR-T treatment. However, given that both patients have higher levels of IL6, it is suggested that severe COVID-19 patients with significantly increased IL6 may also benefit from IL6R monoclonal antibody treatment. Most patients with COVID-19 had normal serum PCT levels on admission, but their serum PCT levels increased when they had a secondary infection[9]. Because patients with low lymphocytes are prone to bacterial infections, dynamic testing for PCT may be helpful in diagnosing patients with bacterial infections.

In conclusion, the counts of CD3^+^, CD4^+^, and CD8^+^ T-cells are significantly reduced in severe patients compare to mild and moderated COVID-19 patients, and the reduction of these T cells is related to the severity of the disease. Both the counts of CD3^+^ and CD4^+^ T-cells can be used as predictive indicators for predicting the severity of COVID-19 patients, and dynamic detection of the T-lymphocyte subsets may help predict the outcome of the patients.

## Data Availability

The data that support the findings of this study are available from the corresponding author upon reasonable request.

## Acknowledgments

This study was supported financially by New Product Development Projects of Yunnan Province (2016BC005), The “Thirteenth Five-Year” Science and Technology Major Project for Prevention and Control of AIDS, Viral Hepatitis and Other Major Infectious Diseases (2017ZX10202101-002-005) and the National Natural Science Foundation of China (8136024).

---

**Table 1.** Baseline Characteristics and Treatments of Patients Infected With SARS-CoV-2

| **Baseline Characteristics** | **Total (N=16)** | **Mild (N=5)** | **Moderate (N=7)** | **Severe (N=4)** |
| --- | --- | --- | --- | --- |
| Age, median (min-max), years | 49 (3-69) | 22 (3-40) | 50 (32-57) | 61 (49-69) |
| Gender |  |  |  |  |
| Men | 9 (56.25%) | 3 (18.75%) | 3 (18.75%) | 3 (18.75%) |
| Women | 7 (43.75%) | 2 (12.50%) | 4 (25.00%) | 1 (6.25%) |
| Residence |  |  |  |  |
| Wuhan | 15 (93.75%) | 5 (31.25%) | 6 (37.50%) | 4 (25.00%) |
| Local | 1 (6.25%) | 0 | 1 (6.25%) | 0 |
| Family clustering cases | 4 (25.00%) | 3 (18.75%) | 1 (6.25%) | 0 |
| Onset of symptom to Hospital admission, median (min-max), days | 6 (1-11) | 2 (1-3) | 7 (5-10) | 7.5 (6-11) |
| Comorbidities | 5 (31.25%) | 1 (6.25%) | 1 (6.25%) | 3 (18.75%) |
| Hypertension | 3 (18.75%) | 1 (6.25%) | 0 | 2 (12.50%) |
| Diabetes | 2 (12.50%) | 0 | 1 (6.25%) | 1 (6.25%) |
| Signs and symptoms |  |  |  |  |
| Fever | 12 (75.00%) | 4 (25.00%) | 5 (31.25%) | 3 (18.75%) |
| <37.3 | 4 (25.00%) | 1 (6.25%) | 2 (12.50%) | 1 (6.25%) |
| 37.3-38 | 8 (50.00%) | 3 (18.75%) | 3 (18.75%) | 2 (12.50%) |
| 38.1-39.0 | 3 (18.75%) | 1 (6.25%) | 1 (6.25%) | 1 (6.25%) |
| >39.0 | 1 (6.25%) | 1 (6.25%) | 0 | 0 |
| Cough | 7 (43.75%) | 1 (6.25%) | 5 (31.25%) | 1 (6.25%) |
| Sputum production | 6 (37.50%) | 1 (6.25%) | 4 (25.00%) | 1 (6.25%) |
| Chest tightness | 2 (12.50%) | 0 | 0 | 2 (12.50%) |
| Shortness of breath | 2 (12.50%) | 0 | 0 | 2 (12.50%) |
| Fatigue | 6 (37.50%) | 0 | 3 (18.75%) | 3 (18.75%) |
| Headache | 1 (6.25%) | 1 (6.25%) | 0 | 0 |
| Pharyngalgia | 1 (6.25%) | 0 | 1 (6.25%) | 0 |
| Body aches | 2 (12.50%) | 0 | 0 | 2 (12.50%) |
| Baseline CT Scan signs |  |  |  |  |
| Normal | 5 (31.25%) | 5 (31.25%) | 0 | 0 |
| Abnormal | 11 (68.75%) | 0 | 7 (43.75%) | 4 (25.00%) |
| **Treatments** |  |  |  |  |
| KALETRA | 14 (87.50%) | 3 (18.75%) | 7 (43.75%) | 4 ((25.00%) |
| IFN-α nebulization inhalation | 16 (100%) | 5 (31.25%) | 7 (43.75%) | 4 (25.00%) |
| Immunoglobulin | 5 (31.25%) | 0 | 1 (6.25%) | 4 (25.00%) |
| Antibiotics | 4 (25.00%) | 0 | 1 (6.25%) | 3 (18.75%) |
| Glucocorticoids | 1 (6.25%) | 0 | 0 | 1(6.25%) |
| **Virus status\*** |  |  |  |  |
| Onset to Virus eradication, rate | 13 (81.25%) | 4 (25.00%) | 6 (37.50%) | 3 (18.75%) |
| Days from Onset to Virus eradication, median (min-max) | 21 (15-43) | 17 (9-27) | 24 (15-43) | 21 (15-22) |

2 \*Virus status is based on the detection of SARS-CoV-2 by RT-qPCR in throat swab samples.

## References

1. Zhu N, Zhang D, Wang W, et al. A Novel Coronavirus from Patients with Pneumonia in China, 2019. N Engl J Med 2020; 382: 727–33.

2. Sahin AR. 2019 Novel Coronavirus (COVID-19) Outbreak: A Review of the Current Literature. Eurasian Journal of Medical Investigation 2020.

3. Lu R, Zhao X, Li J, et al. Genomic characterisation and epidemiology of 2019 novel coronavirus: implications for virus origins and receptor binding. Lancet 2020; 395: 565–74.

4. Naming the coronavirus disease (COVID-2019) and the virus that causes it. Available at: https://www.who.int/emergencies/diseases/novel-coronavirus-2019/technical-guidance/naming-the-coronavirus-disease-(covid-2019)-and-the-virus-that-causes-it.

5. Gorbalenya AE, Baker SC, Baric RS, et al. *Severe acute respiratory syndrome-related coronavirus*: The species and its viruses—a statement of the Coronavirus Study Group. bioRxiv 2020:2020.02.07.937862.

6. Coronavirus disease 2019 (COVID-19) Situation Report—36. Available at: https://www.who.int/docs/default-source/coronaviruse/situation-reports/20200225-sitrep-36-covid-19.pdf?sfvrsn=2791b4e0_2.

7. Chang Lin M, Wei L, et al. Epidemiologic and Clinical Characteristics of Novel Coronavirus Infections Involving 13 Patients Outside Wuhan, China. JAMA 2020.

8. Bai Y, Yao L, Wei T, et al. Presumed Asymptomatic Carrier Transmission of COVID-19. JAMA 2020.

9. Huang C, Wang Y, Li X, et al. Clinical features of patients infected with 2019 novel coronavirus in Wuhan, China. Lancet 2020; 395: 497–506.

10. Wang D, Hu B, Hu C, et al. Clinical Characteristics of 138 Hospitalized Patients With 2019 Novel Coronavirus-Infected Pneumonia in Wuhan, China. JAMA 2020.

11. Xu Z, Shi L, Wang Y, et al. Pathological findings of COVID-19 associated with acute respiratory distress syndrome. Lancet Respir Med 2020.

12. Wong RS, Wu A, To KF, et al. Haematological manifestations in patients with severe acute respiratory syndrome: retrospective analysis. BMJ 2003; 326: 1358–62.

13. Chu H, Zhou J, Wong BH, et al. Middle East Respiratory Syndrome Coronavirus Efficiently Infects Human Primary T Lymphocytes and Activates the Extrinsic and Intrinsic Apoptosis Pathways. J Infect Dis 2016; 213: 904–14.

14. Gu J, Gong E, Zhang B, et al. Multiple organ infection and the pathogenesis of SARS. J Exp Med 2005; 202: 415–24.

15. Panesar NS, Lam CW, Chan MH, Wong CK, Sung JJ. Lymphopenia and neutrophilia in SARS are related to the prevailing serum cortisol. Eur J Clin Invest 2004; 34: 382–4.

16. Drewry AM, Samra N, Skrupky LP, Fuller BM, Compton SM, Hotchkiss RS. Persistent lymphopenia after diagnosis of sepsis predicts mortality. Shock 2014; 42: 383–91.

17. Gomersall CD. Pro/con clinical debate: steroids are a key component in the treatment of SARS. Pro: Yes, steroids are a key component of the treatment regimen for SARS. Crit Care 2004; 8: 105–7.

18. Liu J, Liu Y, Xiang P, et al. Neutrophil-to-Lymphocyte Ratio Predicts Severe Illness Patients with 2019 Novel Coronavirus in the Early Stage. medRxiv 2020:2020.02.10.20021584.

19. Liu J, Li S, Liu J, et al. Longitudinal characteristics of lymphocyte responses and cytokine profiles in the peripheral blood of SARS-CoV-2 infected patients. medRxiv 2020:2020.02.16.20023671.

20. de Wolf F, Spijkerman I, Schellekens PT, et al. AIDS prognosis based on HIV-1 RNA, CD4^+^ T-cell count and function: markers with reciprocal predictive value over time after seroconversion. AIDS 1997; 11: 1799–806.

21. Mudd JC, Lederman MM. CD8 T cell persistence in treated HIV infection. Curr Opin HIV AIDS 2014; 9: 500–5.

22. Plesko M, Suvada J, Makohusova M, et al. The role of CRP, PCT, IL-6 and presepsin in early diagnosis of bacterial infectious complications in paediatric haemato-oncological patients. Neoplasma 2016; 63: 752–60.

23. Chew KS. What’s new in Emergencies Trauma and Shock? C-reactive protein as a potential clinical biomarker for influenza infection: More questions than answers. J Emerg Trauma Shock 2012; 5: 115–7.

24. Schuetz P, Albrich W, Mueller B. Procalcitonin for diagnosis of infection and guide to antibiotic decisions: past, present and future. BMC Med 2011; 9: 107.

25. Jawa RS, Anillo S, Huntoon K, Baumann H, Kulaylat M. Interleukin-6 in surgery, trauma, and critical care part II: clinical implications. J Intensive Care Med 2011; 26: 73–87.

26. Chen H, Guo J, Wang C, et al. Clinical characteristics and intrauterine vertical transmission potential of COVID-19 infection in nine pregnant women: a retrospective review of medical records. The Lancet 2020.

27. Chan JF, Yuan S, Kok KH, et al. A familial cluster of pneumonia associated with the 2019 novel coronavirus indicating person-to-person transmission: a study of a family cluster. Lancet 2020; 395: 514–23.

28. Zhang Y, Li J, Zhan Y, et al. Analysis of serum cytokines in patients with severe acute respiratory syndrome. Infect Immun 2004; 72: 4410–5.

29. Shimabukuro-Vornhagen A, Godel P, Subklewe M, et al. Cytokine release syndrome. J Immunother Cancer 2018; 6: 56.

30. Maude SL, Barrett D, Teachey DT, Grupp SA. Managing cytokine release syndrome associated with novel T cell-engaging therapies. Cancer J 2014; 20: 119–22.

